# Maternal & Infant Health Benefits of a Nicotine Product Standard in the United States

**DOI:** 10.1101/2025.09.11.25335605

**Authors:** Atalay Demiray, Sarah Skolnick, Jamie Tam

**Affiliations:** Department of Health Policy and Management Yale School of Public Health New Haven, CT, United States; Department of Health Behavior, Society, and Policy Rutgers School of Public Health Rutgers Institute on Nicotine and Tobacco Studies

## Abstract

**Importance:** Cigarette smoking during pregnancy increases the risks of miscarriage, ectopic pregnancy, placental complications, hypertensive disorders of pregnancy, and infant mortality. Reducing smoking in pregnancy remains a pressing public health priority.

**Objective:** To project the impact of a proposed nicotine product standard on maternal complications and infant mortality using the Smoking, E-cigarette use, and Pregnancy (SEP) microsimulation model.

**Design, Settings, and Participants:** Individual-level, annual‐cycle microsimulation of U.S. females of reproductive age that tracks smoking, vaping, pregnancy, and pregnancy outcomes was constructed. Model inputs were drawn from national surveillance and vital statistics, including National Health Interview Survey (NHIS) for general population, Behavioral Risk Factor Surveillance System (BRFSS) (2016–2023) for smoking/vaping among pregnant women and NVSS Natality/Linked Birth–Infant Death files for late-pregnancy morbidities and infant mortality. Policy effects on tobacco and e-cig usage transitions are based on FDA’s expert-elicitation and used to simulate outcomes under status quo vs. the new policy from 2027-2100.

**Exposure:** Smoking and vaping.

**Main Outcome(s):** Maternal outcomes during pregnancy (ectopic pregnancy, miscarriage, placenta previa, placental abruption, hypertensive disorders of pregnancy/pre-eclampsia, eclampsia), infant mortality, pregnancy-related costs, and maternal Quality Adjusted Life Years (QALYs).

**Results:** Under the nicotine product standard (policy start 2027), smoking in pregnancy falls sharply from 6.0% in 2027 to 1.2% by 2040. These behavioral shifts translate into large perinatal gains through 2100: approximately 167,000 ectopic pregnancies, 950,000 miscarriages, 15,000 placenta previa, 62,000 placental abruptions, 167,000 hypertensive disorders of pregnancy/pre-eclampsia, 9,000 eclampsia cases, and 64,000 infant deaths are averted cumulatively. Maternal health improves as well, with 103,000 pregnancy QALYs gained. Health system spending falls despite conservative costing, with $4.9 billion in pregnancy-related medical costs avoided. Across uncertainty bounds, direction and magnitude of benefit remain favorable for all maternal morbidities and infant deaths, indicating that new policy yields substantial and durable health gains alongside meaningful cost offsets.

**Conclusions:** A proposed nicotine product standard is projected to improve maternal and infant outcomes and yield sizable pregnancy-related health gains and cost offsets. The SEP model complements prior tobacco policy evaluation frameworks while focusing on maternal and infant health.

## Introduction

Cigarette smoking during pregnancy is a well-established risk factor for adverse maternal and infant outcomes, including ectopic pregnancy, spontaneous miscarriage, placental complications, preterm delivery, and infant mortality.^1,2^ Maternal smoking roughly doubles the odds of placenta previa and placental abruption and increases ectopic pregnancy risk by ∼1.7-fold.^3-5^ Maternal smoking also leads to reduced fetal growth and is associated with higher neonatal and infant mortality rates.^2^ These pregnancy-related harms add to the overall health burden of tobacco use and underscore the importance of effective smoking cessation, especially in women of reproductive age. Yet 44% of those who smoked before pregnancy continue smoking during pregnancy.^6^ These are abysmal statistics that underscore the difficulty of smoking cessation and the ongoing burden of cigarette smoking on maternal and infant health. At the same time, the use of e-cigarettes has risen in the general population, including among young women.^7^ E-cigarette use is now a common quitting method reported by adults who smoke, ranking as high as nicotine replacement therapy in recent years.^8^ The implications of e-cigarette use during pregnancy remain uncertain.^9^

Bold policy intervention could accelerate improvements to pregnancy outcomes across the population. The Food and Drug Administration (FDA) has proposed a rule that would reduce nicotine levels in cigarettes to minimally addictive levels, also known as a ‘nicotine product standard.’^10^ Because continued smoking is driven by nicotine addiction, this rule could dramatically increase rates of successful smoking cessation, thereby reducing the burden of tobacco on maternal and infant health outcomes.^11^ Randomized clinical trials show that women of reproductive age who are randomized to receive very-low-nicotine cigarettes reduce their smoking compared to those who receive normal-nicotine cigarettes.^12^ Prior studies have modeled the long-term impact of a nicotine product standard on overall mortality and on mental health outcomes.^13,14^ The potential long-term impact of a nicotine standard for maternal and infant health outcomes has never been evaluated. We developed the Smoking, E-cigarette Use, and Pregnancy (SEP) microsimulation model, an individual-level simulation of U.S. women that incorporates smoking, e-cigarette use, and pregnancy. This model integrates tobacco-use dynamics with pregnancy outcomes and can inform policymakers of the potential benefits (or unintended consequences) of such interventions. The SEP model simulates cohorts of women over time, tracking their smoking/vaping status, pregnancies, and pregnancy outcomes under varying nicotine product standard scenarios. By capturing the interplay between smoking behavior and pregnancy, the model estimates the population-level impact of the standard on pregnancy outcomes (e.g., miscarriages, complications, infant deaths) as well as related costs and quality of life.

## Methods

### Model overview

We developed the SEP microsimulation, an individual-level, annual-cycle state-transition model of U.S. females of reproductive age, 15-49. SEP tracks three concurrent dimensions: smoking (never/current/former), vaping (never/current/former), and pregnancy status (pregnant/not pregnant) while updating them each year to capture conception, gestation, pregnancy outcomes, and postpartum relapse/cessation dynamics. We simulated 10,000 women for each birth cohort from 1900 to 2100, totaling 1 million in the modeled population. We used an annual cycle for computational simplicity and alignment to annual data inputs and pregnancy is approximated as occurring within the cycle of conception. Figure 1 depicts the 19 health states and transitions, the pregnancy module, and the infant outcome decision tree. We simulated a federal nicotine product standard beginning in 2027 (consistent with the FDA’s proposed rule) and assessed health and economic impacts through 2100, reporting smoking and vaping prevalence among pregnant people, infant deaths averted, and maternal pregnancy QALYs gained.

**Figure 1.**
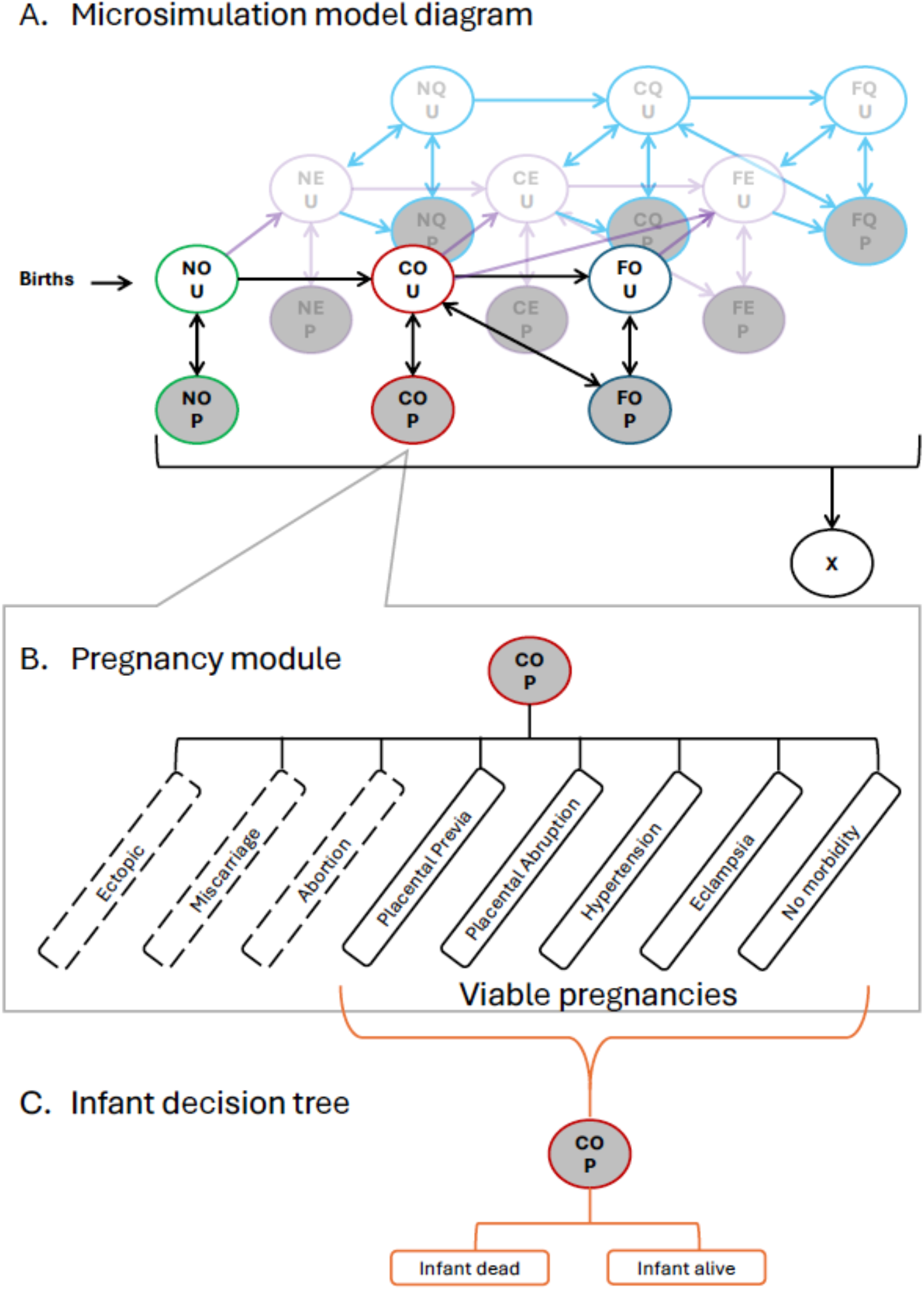
Overview of the product use and pregnancy status transitions in the Smoking, E-cigarette use, and Pregnancy (SEP) Model. **Note:** Self-loops (“stay” transitions) are omitted from the diagram. In each annual cycle, individuals may remain in their current state; for every state, the probabilities of all depicted outgoing transitions plus the (omitted) stay probability sum to 1.

### Data sources & parameterization

Transitions between smoking states were based on the CISNET Lung Working Group estimates of annual smoking initiation and cessation.^15^ SEP microsimulation model captures pregnancy-specific behavior: smokers have elevated cessation when transitioning to pregnancy state with relapse risk in the year after delivery, while initiation during pregnancy is assumed negligible. Evidence on pregnancy-specific vaping is sparse; in the base case we do not impose pregnancy-specific vaping transitions beyond those in the general population.^9^

We also used mortality probabilities for the general population, which vary by smoking status and years since quit, from the CISNET Lung Working Group.^15^ Due to the low number of maternal deaths, we do not model additional mortality associated with pregnancy beyond mortality for the general population.

We calibrated baseline smoking and vaping prevalence among general population by age using NHIS (2014–2022) and used BRFSS (2016–2023) to target smoking/vaping prevalence among pregnant women.^16,17^ While NHIS is more representative for general population, BRFSS includes larger pregnancy sample, hence more reliable for pregnant population. Natality data (2013–2023) provide maternal smoking by trimester and pregnancy complications while NSFG (2021–2023) refines early-loss probabilities including miscarriage, ectopic pregnancy by age and smoking.^18,19^

### Pregnancy Module and Pregnancy Outcomes

In each cycle, non-pregnant women can conceive based on age-specific pregnancy probabilities derived from vital statistics and survey data. We also explicitly model closely spaced pregnancies: for second pregnancies, we assume 16.2% occur within 12 months (short interpregnancy interval), so a small number of individuals remain in the pregnancy state in consecutive cycles due to conception <12 months postpartum, based on prior estimates.^20^

Early outcomes (miscarriage, ectopic pregnancy, induced abortion) are parameterized using NSFG and stratified by maternal age and smoking status. For pregnancies reaching ≥20 weeks, late-pregnancy outcomes (placenta previa, placental abruption, gestational hypertension/pre-eclampsia, eclampsia) and infant mortality are sourced from NVSS Natality and the CDC Linked Birth–Infant Death datasets, using smoking-stratified risks.^18,21^ Where NVSS stopped collecting placenta previa/abruption (post-2005), we carry forward stable 1995–2005 estimates and smoking differentials. Infant mortality probabilities per 1,000 live births are ∼7.5 in smokers vs ∼5.0 in never-smokers (former smokers intermediate) and are held constant after 2023.^21^

### Nicotine Product Standard

We simulate a federal nicotine product standard beginning in 2027 and run through 2100 using the FDA expert elicitation.^22^ Policy effects were applied to initiation, cessation, dual use, switching, and vaping initiation among those deterred from smoking. We also examine optimistic/pessimistic bounds using the expert elicitation’s 5th and 95th percentiles.^23^ All outcomes are reported relative to a status-quo scenario with unchanged tobacco transitions.

### Structural assumptions

Key structural assumptions are (i) static near-term fertility projections; (ii) one-year pregnancy cycle approximation; (iii) former smokers who quit early in pregnancy carry baseline risks for most outcomes; (iv) insufficient evidence to assign pregnancy-specific e-cigarette effects (treated as neutral in base case); and (v) no additional pregnancy-specific hazard for all-cause maternal mortality (given its low absolute risk).

### Outcomes

We report cumulative differences (policy vs. status quo): counts of pregnancy outcomes, pregnancy-related medical costs, and maternal QALYs. Infant QALYs are not tallied as infants are not explicitly followed in SEP, and infant outcomes enter through maternal costs and infant deaths. Costs are drawn from published sources and inflated to 2023 USD (e.g., ectopic ≈$20,000, miscarriage ≈$697, previa ≈$17,500, abruption ≈$43,000, hypertensive disorders ≈$13,000, eclampsia ≈$9,000, infant death ≈$77,000).^24-26^ We simulate large cohorts to obtain stable estimates of pregnancy events, costs, and QALYs under policy vs status quo. Calibration and validation targets were the observed as prevalences by age and smoking status while uncertainty is characterized using the FDA policy-effect bounds.^22,23^

## Results

### Smoking trajectories in pregnancy

With a nicotine product standard beginning in 2027, projected smoking in pregnancy falls rapidly (Figure 2). Pregnant-smoker prevalence declines from 6.0% in 2027 to 1.2% by 2040 under the product standard. Without this standard, smoking prevalence stabilizes at ∼3% from 2040 onward. Following recent trends, vaping is expected to continue increasing among pregnant women. Initially, vaping prevalence under the product standard even surpasses that of the status quo, due to individuals deterred from smoking who instead take up vaping. After this initial increase, the product standard moderately reduces levels of vaping compared to status quo: in 2100, there is an 8% vaping prevalence under status quo compared to 7% under the product standard. From 2027 to 2100, dual use remains quite low at ∼1% under status quo and ∼0% under a nicotine product standard.

**Figure 2.**
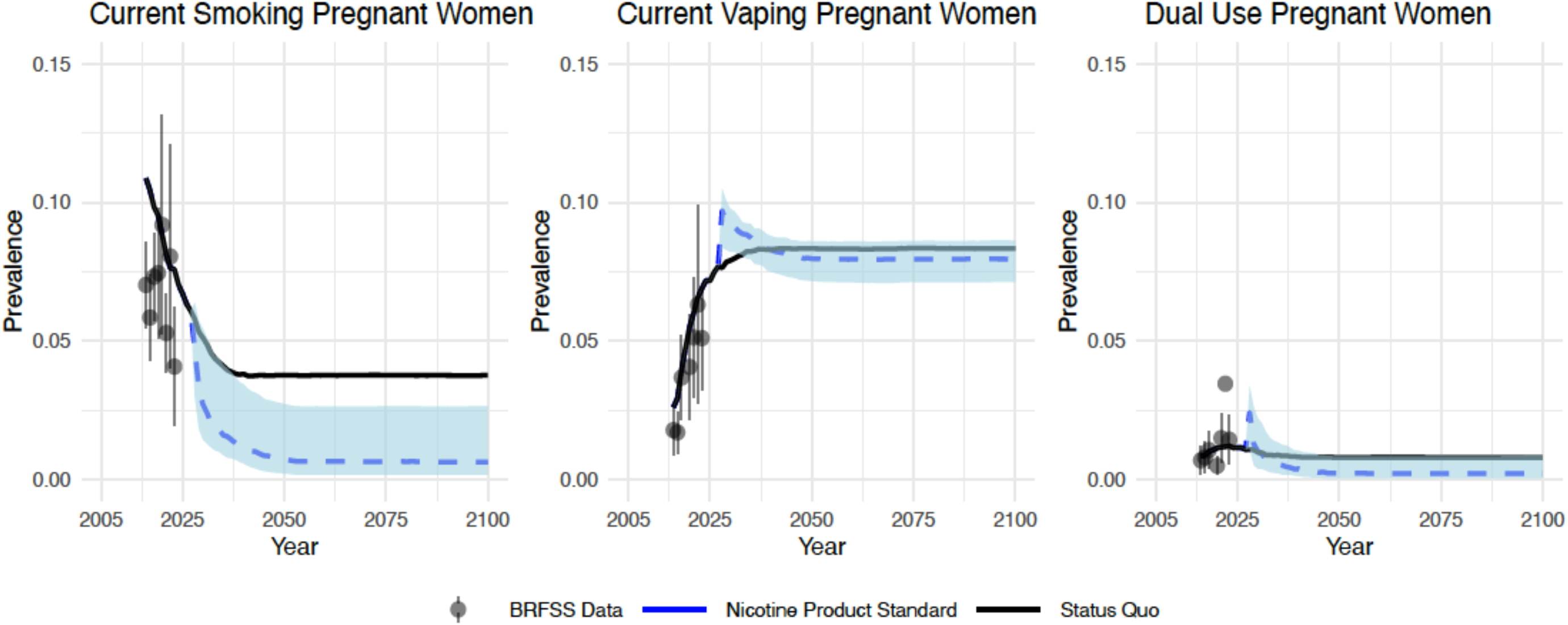
Actual and projected Smoking and Vaping Prevalence among Pregnant Women 2015-2100

### Maternal and infant outcomes

Reductions in smoking during pregnancy translate into large perinatal gains under a nicotine product standard, with outcomes consistently favorable across pessimistic–optimistic policy bounds (Table 2). Notably, 167,000 (97,000-228,000) ectopic pregnancies, 200,000 (148,000 - 306,000) cases of gestational hypertension and 9,000 (5,000-14,000) cases of eclampsia were averted from 2027 to 2100 under the nicotine product standard. The SEP model also projects 15,000 (5,000–19,000) placenta previa events averted and 62,000 (19,000–75,000) fewer placental abruptions, reducing risks of antepartum bleeding and emergent delivery. Importantly, these upstream improvements cascade to neonatal outcomes: we estimate 64,000 infant deaths averted (27,000–82,000) over 2027–2100, reflecting fewer catastrophic obstetric events due to lower exposure to in-utero tobacco smoke.

**Table 1.** Model inputs and primary data sources

| Domain | Key inputs | Primary source(s) |
| --- | --- | --- |
| Smoking/vaping transitions (baseline) | Initiation, cessation, dual-use probabilities; calibration to U.S. surveillance | Prior transition frameworks; NHIS/BRFSS |
| Policy effects (nicotine product standard) | Changes to initiation, cessation, dual use, switching; start year 2027 | FDA expert-elicitation |
| Pregnancy incidence | Age-specific conception/pregnancy probabilities; near-term static trend assumption | NVSS Natality |
| Early losses | Miscarriage, ectopic, induced abortion by age and smoking status | NSFG |
| Late pregnancy outcomes | Placenta previa, placental abruption, hypertensive disorders, eclampsia by smoking status | NVSS Natality |
| Infant mortality | Linked Birth–Infant Death (IMR) by maternal smoking status | CDC linked files |
| Costs (2023 USD) | Ectopic (\$20k), miscarriage (\$697), previa (\$17.5k), abruption (\$43.25k), HPE (\$13k), eclampsia (\$9k), infant death (~\$77k) | Various Literature |
| Utilities | Per-pregnancy QALY decrements (disutility) | Various Literature |

**Table 2.** Cumulative Pregnancy Outcomes (Number of events averted, QALYs gained, and costs averted)

| **Year** | **Healthy**<br>**Pregnancies**<br>**Gained** | **Ectopic**<br>**Pregnancies**<br>**averted** | **Placenta**<br>**Previa**<br>**averted** | **Placental**<br>**Abruptio**<br>**Averted** | **Hypertensi**<br>**on Averted** | **Eclampsia**<br>**Averted** | **Infant**<br>**Deaths**<br>**Averted** | **Medical**<br>**cost savings**<br>**(billions)** | **QALYs**<br>**gained** |
| --- | --- | --- | --- | --- | --- | --- | --- | --- | --- |
| **2027** | **0** | **0** | **0** | **0** | **0** | **0** | **0** | **0** | **0** |
| **2040** | **84**<br>**(136 - 11)** | **7**<br>**(2 - 11)** | **2**<br>**(-1 - 3)** | **9**<br>**(-1 - 13)** | **-15**<br>**(-12 - -5)** | **0**<br>**(0 - 0)** | **6**<br>**(0 - 8)** | **0.6**<br>**(-0.14 - 1.06)** | **74**<br>**(108 - 23)** |
| **2060** | **34**<br>**(-131 - 134)** | **57**<br>**(32 - 79)** | **6**<br>**(1 - 8)** | **27**<br>**(5 - 34)** | **46**<br>**(41 - 80)** | **2**<br>**(1 - 3)** | **24**<br>**(8 - 32)** | **2.77**<br>**(1.01 - 3.96)** | **89**<br>**(130 - 9)** |
| **2080** | **-27**<br>**(-284 - 122)** | **113**<br>**(65 - 155)** | **11**<br>**(3 - 14)** | **45**<br>**(12 - 55)** | **124**<br>**(95 - 195)** | **6**<br>**(3 - 9)** | **44**<br>**(18 - 58)** | **4.15**<br>**(1.77 - 5.79)** | **98**<br>**(142 - 3)** |
| **2100** | **-88**<br>**(-436 - 113)** | **167**<br>**(97 - 228)** | **15**<br>**(5 - 19)** | **62**<br>**(19 - 75)** | **200**<br>**(148 - 306)** | **9**<br>**(5 - 14)** | **64**<br>**(27 - 82)** | **4.9**<br>**(2.17 - 6.77)** | **103**<br>**(-1 - 148)** |

**Note: Outcomes are cumulative with optimistic and pessimistic scenarios provided in parentheses. Medical cost savings is in billions and all other values are presented as is. \*All values are in 1000s except for Medical cost savings which is in billions.**

### Costs and QALYs

These clinical gains yield meaningful economic and quality-of-life benefits. By 2100, the SEP model estimates ∼$4.9 billion ($2.17–$6.77 billion) in pregnancy-related medical cost savings with the largest contributions coming from avoided hypertensive disorders, placental abruption, and the high acute costs associated with infant death. Maternal health-related quality of life also improves, with ∼103,000 pregnancy QALYs gained (−1,000 to 148,000). Across pessimistic– optimistic policy bounds, effects remain uniformly favorable in direction and clinically important in magnitude, indicating that near-elimination of smoking in pregnancy produces durable perinatal gains alongside substantial cost offsets (Table 2).

## Discussion

This is the first study to quantify the impact of a nicotine product standard on maternal and infant health outcomes by integrating smoking–vaping dynamics with conception, pregnancy morbidities, and infant survival. In the SEP model, a new nicotine standard drives steep declines in smoking during pregnancy and is associated with large reductions in early losses (miscarriage, ectopic pregnancy), placental complications, hypertensive disorders of pregnancy and eclampsia, and infant deaths.^1-5^ Maternal health improves (≈159,000 pregnancy QALYs gained; range 42,000–268,000) while pregnancy-related medical costs decline (≈$4.81B averted; range $1.62B– $6.80B).

The direction and magnitude of benefits are consistent with well-established links between cigarette smoking and adverse pregnancy outcomes as well as infant mortality. By reducing addiction and facilitating smoking cessation, the nicotine standard lowers exposure to upstream health risk, thereby improving downstream perinatal outcomes.^2,11^ Our findings complement recent nicotine-standard modeling conducted at the general-population level (and in priority subgroups such as people with major depression), which projects large declines in smoking and substantial health and economic gains under a 2027–2100 policy horizon.^13,22^ By explicitly linking tobacco transitions to conception, pregnancy outcomes, and infant survival, SEP extends that policy lens to maternal and infant health, a domain not directly evaluated in prior studies.

A nicotine product standard is likely to generate durable perinatal benefits and pregnancy-year cost offsets even under conservative modeling choices (e.g., excluding infant QALYs and long-term maternal sequelae). These gains are additive to population-wide mortality and productivity improvements anticipated under the policy, strengthening the case for timely implementation and for integrating the rule with pregnancy-specific cessation supports (e.g., prenatal counseling, postpartum relapse prevention).

Methodologically, SEP (i) couples smoking/vaping trajectories to pregnancy incidence and outcomes; (ii) calibrates pregnant-subgroup smoking/vaping to BRFSS 2016–2023; and (iii) draws maternal and infant outcomes from NVSS Natality and Linked Birth–Infant Death files, using smoking-stratified risks. These modeling choices allow valid replication of observed pregnancy morbidities while maintaining policy relevance. The policy effects applied in the model leverages FDA’s expert-elicitation approach used in prior nicotine-standard evaluations.^22,23^ These represent the best available data on the likely impacts of a nicotine standard given that no country in the world has implemented one yet.

Several simplifying assumptions warrant caution in interpretation. First, SEP uses annual cycles and approximates each pregnancy within the year of conception, concentrating costs and utilities in a single year and precluding multiple pregnancies in the same year. Second, due to limited evidence, the base case assumes no direct effect of vaping on pregnancy outcomes.^9^ Third, infant outcomes are not followed as distinct entities beyond the birth year; infant/child QALYs are not counted and “saved” infants are not added to future cohorts. Fourth, placenta previa and abruption inputs rely on historical vital statistics reporting and we assume stable rates and smoking differentials that may not fully reflect contemporary obstetric practice. Fifth, utility decrements emphasize acute maternal burden and omit longer-term psychological sequelae after pregnancy loss. Sixth, early-pregnancy losses draw on surveys subject to underreporting and SEP currently applies abortion probabilities uniformly across smoking categories and assumes no smoking-specific fertility differential. Finally, as in prior work, policy effects are based on expert elicitation rather than real world evidence; illicit markets and non-cigarette combustibles are not modeled, which could attenuate the direction of benefits.

Perinatal benefits and pregnancy-year cost offsets add an important dimension to the nicotine-standard evidence base. Implementation could be complemented by targeted prenatal cessation support and postpartum relapse prevention to maximize maternal–infant gains. Future model extensions might include trimester-level resolution, incorporation of infant/child QALYs and longer-term maternal outcomes, and stratified projections by education and other sociodemographic factors to assess equity impacts. As regulatory timelines evolve, states or countries considering nicotine-reduction policies can use SEP’s perinatal projections as one component of a broader policy assessment that includes surveillance for product substitution and real-world effects on pregnancy and neonatal outcomes.

## Conclusions

A nicotine product standard is projected to yield substantial, durable improvements in maternal and infant health alongside meaningful pregnancy-year cost offsets. By centering maternal and infant health, SEP complements prior nicotine-standard evaluations and underscores the maternal– infant health value of strong nicotine regulation.

## Data Availability

All data used in this study are publicly available online from the US Centers for Disease Control and Prevention (CDC) and the National Center for Health Statistics (NCHS). Specifically: adult tobacco use data from NHIS, BRFSS questionnaires for constructing smoking/vaping indicators, NVSS Natality Public Use documentation, and NSFG survey files and documentation. Direct access links:
NHIS Adult Tobacco Use overview: https://archive.cdc.gov/www_cdc_gov/nchs/nhis/tobacco/tobacco_overview.htm BRFSS Questionnaires: https://www.cdc.gov/brfss/questionnaires/index.htm NVSS Natality Public Use File (User Guide 2023): https://ftp.cdc.gov/pub/health_statistics/nchs/dataset_documentation/DVS/natality/UserGuide2023.pdf NSFG Homepage: https://www.cdc.gov/nchs/nsfg/index.htm These resources are open-access and contain only deidentified public-use data. No special permissions or data-sharing agreements are required to obtain them.

https://ftp.cdc.gov/pub/health_statistics/nchs/dataset_documentation/DVS/natality/UserGuide2023.pdf

https://www.cdc.gov/brfss/questionnaires/index.htm

https://archive.cdc.gov/www_cdc_gov/nchs/nhis/tobacco/tobacco_overview.htm

## References

1. American College of Obstetricians and Gynecologists. Tobacco and nicotine cessation during pregnancy. Committee Opinion No. 807. Obstet Gynecol. 2020;135(5):e221–e229. doi:10.1097/AOG.0000000000003822

2. US Department of Health and Human Services. The Health Consequences of Smoking—50 Years of Progress: A Report of the Surgeon General. Atlanta, GA: US Dept of Health and Human Services, Centers for Disease Control and Prevention; 2014. Accessed September 10, 2025. https://www.ncbi.nlm.nih.gov/books/NBK179276/

3. Shobeiri F, Jenabi E. Smoking and placenta previa: a meta-analysis. J Matern Fetal Neonatal Med. 2017;30(24):2985–2990. doi:10.1080/14767058.2016.1271405

4. Ananth CV, Smulian JC, Vintzileos AM. Incidence of placental abruption in relation to cigarette smoking and hypertensive disorders during pregnancy: a meta-analysis of observational studies. Obstet Gynecol. 1999;93(4):622–628. doi:10.1016/S0029-7844(98)00408-6

5. Horne AW, Brown JK, Nio-Kobayashi J, et al. The association between smoking and ectopic pregnancy: why nicotine is BAD for your Fallopian tube. PLoS One. 2014;9(2):e89400. doi:10.1371/journal.pone.0089400

6. Kipling L, Bombard J, Wang X, Cox S. Cigarette smoking among pregnant women during the perinatal period: prevalence and health care provider inquiries—Pregnancy Risk Assessment Monitoring System, United States, 2021. MMWR Morb Mortal Wkly Rep. 2024;73(17):393–398. doi:10.15585/mmwr.mm7317a2

7. Vahratian A, Briones EM, Jamal A, Marynak KL. Electronic cigarette use among adults in the United States, 2019–2023. NCHS Data Brief. 2025;(524):1–8. Accessed September 10, 2025. https://www.cdc.gov/nchs/products/databriefs/db524.htm

8. Gravely S, Cummings KM, Hammond D, et al. Self-reported quit aids and assistance used by smokers at their most recent quit attempt: findings from the 2020 International Tobacco Control Four Country Smoking and Vaping Survey. Nicotine Tob Res. 2021;23(10):1699–1707. doi:10.1093/ntr/ntab068.

9. Ussher M, Fleming J, Brose L. Vaping during pregnancy: a systematic review of health outcomes. BMC Pregnancy Childbirth. 2024;24:435. doi:10.1186/s12884-024-06633-6

10. Food and Drug Administration. Tobacco product standard for nicotine yield of cigarettes and certain other combusted tobacco products. Published January 16, 2025. Accessed September 10, 2025. https://www.federalregister.gov/documents/2025/01/16/2025-00397/tobacco-product-standard-for-nicotine-yield-of-cigarettes-and-certain-other-combusted-tobacco

11. Food and Drug Administration. Final Summary Report: The Science of a Nicotine Standard for Combusted Tobacco Products. Published January 15, 2025. Accessed September 10, 2025. https://www.fda.gov/media/185051/download

12. Higgins ST, Sigmon SC, Tidey JW, et al. Reduced nicotine cigarettes and e-cigarettes in high-risk populations: 3 randomized clinical trials. JAMA Netw Open. 2024;7(9):e2431731. doi:10.1001/jamanetworkopen.2024.31731

13. Skolnick S, Brouwer A, Cheng C, Tam J. Health, equity, and economic impacts of a nicotine product standard in the United States for people with and without major depression. medRxiv. Posted July 11, 2025. doi:10.1101/2025.07.10.25331302

14. Apelberg BJ, Feirman SP, Salazar E, et al. Potential public health effects of reducing nicotine levels in cigarettes in the United States. N Engl J Med. 2018;378(18):1725–1733. doi:10.1056/nejmsr1714617.

15. Jeon J, Meza R, Krapcho M, Clarke LD, Byrne J, Levy DT. Actual and Counterfactual Smoking Prevalence Rates in the US population via Micro-simulation. Risk Anal. 2012;32(Suppl 1):S51. doi:10.1111/J.1539-6924.2011.01775.X

16. National Center for Health Statistics. NHIS—Adult Tobacco Use: Overview of Topics. Accessed September 11, 2025. https://archive.cdc.gov/www_cdc_gov/nchs/nhis/tobacco/tobacco_overview.htm

17. Centers for Disease Control and Prevention. Behavioral Risk Factor Surveillance System (BRFSS) Questionnaires, 2016–2023. Accessed September 11, 2025. https://www.cdc.gov/brfss/questionnaires/index.htm

18. National Center for Health Statistics. User Guide to the 2023 Natality Public Use File. Hyattsville, MD: CDC/NCHS; 2024. Accessed September 11, 2025. https://ftp.cdc.gov/pub/health_statistics/nchs/dataset_documentation/DVS/natality/UserGuide2023.pdf

19. National Center for Health Statistics. National Survey of Family Growth (NSFG) Homepage. Accessed September 11, 2025. https://www.cdc.gov/nchs/nsfg/index.htm

20. Admon LK, MacCallum-Bridges C, Daw JR. Trends in short interpregnancy interval births in the United States, 2016-2022. Obstet Gynecol. 2025;145(1):82–90. doi:10.1097/AOG.0000000000005784.

21. National Center for Health Statistics. Linked Birth/Infant Death data. Accessed September 11, 2025. https://www.cdc.gov/nchs/nvss/linked-birth-infant-death.htm

22. U.S. Food and Drug Administration. Population Health Model Code and Inputs (Nicotine Product Standard). Published 2025. Accessed September 11, 2025. https://www.fda.gov/media/185147/download

23. U.S. Food and Drug Administration. FDA’s Response to External Peer Review on the Methodological Approach to Modeling the Potential Impact of a Nicotine Product Standard on Tobacco Use, Morbidity, and Mortality in the U.S.Published January 15, 2025. Accessed September 11, 2025. https://www.fda.gov/media/185055/download

24. Stevens W, Shih T, Incerti D, et al. Short-term costs of preeclampsia to the United States health care system. Am J Obstet Gynecol. 2017;217(3):237–248.e16. doi:10.1016/j.ajog.2017.06.060

25. Health System Tracker (KFF-Peterson). Health costs associated with pregnancy, childbirth, and postpartum care. Updated 2023. Accessed September 11, 2025. https://www.healthsystemtracker.org/brief/health-costs-associated-with-pregnancy-childbirth-and-postpartum-care/

26. Nagendra D, Schreiber CA, Sonalkar S, et al. Medical management of early pregnancy loss is cost-effective compared with office uterine aspiration. Am J Obstet Gynecol. 2022;227(2):240–248.e3. doi:10.1016/j.ajog.2022.06.030

